# Assessing the value and knowledge gains from an online tick identification and tick-borne disease management course for the Southeastern United States

**DOI:** 10.1101/2024.03.13.24304232

**Authors:** Catherine A. Lippi, Holly D. Gaff, Alexis L. White, Sadie J. Ryan

## Abstract

**Background:** Tick-borne diseases are a growing public health threat in the United States. Despite the prevalence and rising burden of tick-borne diseases, there are major gaps in baseline knowledge and surveillance efforts for tick vectors, even among vector control districts and public health agencies. To address this issue, an online tick training course (OTTC) was developed through the Southeastern Center of Excellence in Vector-Borne Diseases (SECOEVBD) to provide a comprehensive knowledge base on ticks, tick-borne diseases, and their management.

**Methods:** The OTTC consisted of training modules covering topics including tick biology, tick identification, tick-borne diseases, and public health, personal tick safety, and tick surveillance. The course was largely promoted to vector control specialists and public health employees throughout the Southeastern US. We collected assessment and survey data on participants to gauge learning outcomes, perceptions of the utility of knowledge gained, and barriers and facilitators to applying the knowledge in the field.

**Results:** The OTTC was successful in increasing participants’ baseline knowledge across all course subject areas, with the average score on assessment increasing from 62.6% (pre-course) to 86.7% (post-course). More than half of participants (63.6%) indicated that they would definitely use information from the course in their work. Barriers to using information identified in the delayed assessment included lack of opportunities to apply skills (18.5%) and the need for additional specialized training beyond what the OTTC currently offers (18.5%), while the main facilitator (70.4%) for applying knowledge was having opportunities at work, such as an existing tick surveillance program.

**Conclusions:** Overall, this OTTC demonstrated capacity to improve knowledge in a necessary and underserved public health field, and more than half of participants use or plan to use the information in their work. The geographic reach of this online resource was much larger than simply for the Southeastern region for which it was designed, suggesting a much broader need for this resource. Understanding the utility and penetrance of training programs such as these is important for refining materials and assessing optimal targets for training.

## Background

Tick-borne diseases (TBDs) are increasingly recognized as a growing public health threat in the United States, where reported cases of TBDs like Lyme disease and spotted fever group rickettsiosis (SFGR) have sharply increased in recent decades [1–3]. Further, recent range expansions and introductions of medically important tick vectors, along with the emergence and discovery of novel tick-borne pathogens, underscore the increasing public health burden of TBDs [4, 5]. Despite the increased attention from public health agencies in recent years, TBDs in the United States likely remain substantially underreported [6, 7]. Underreporting can arise due to a variety of factors, often in combination. Constraints such as limitations in diagnostic tools or surveillance capabilities, non-specific clinical presentations, or asymptomatic cases, can stymie case reporting and diagnostics at the point of care. Despite the serious health implications of TBDs, cases may go undiagnosed by clinicians unfamiliar with TBD risk factors and case pathologies. Knowledge gaps among public health professionals can also pose a major obstacle for both tick and TBD surveillance and prevention efforts. Establishing baseline TBD knowledge in frontline workers is all the more crucial now, given the increasing burden and geographic range of many notable TBDs [8].

In the Southeastern United States, TBDs pose substantial risk to public health; diseases like SFGR, ehrlichiosis, and increasingly Lyme disease [9] are reported. Medically important ticks in this region include the lone star tick (*Amblyomma americanum*), Gulf Coast tick (*Amblyomma maculatum*), American dog tick (*Dermacentor variabilis*), brown dog tick (*Rhipicephalus sanguineus*), and the blacklegged tick (*Ixodes scapularis*) [10]. Ticks are also implicated in the spread of diseases with unknown etiology, like southern tick-associated rash illness (STARI) [11, 12], or trigger alpha-gal syndrome (AGS), an allergic response that can develop from exposure to tick bites [13]. Thus, the regional needs of the Southeastern US, in terms of both TBD and tick identification knowledge may differ from e.g. the Northeastern US, which has a necessary focus on diseases caused by *I. scapularis* associated pathogens (e.g., Lyme disease).

Although there have been major advances in efforts to estimate risk and control tick populations, preventing tick bites remains the primary means of reducing exposure to tick-borne pathogens. Identification of risky activities and the adoption of appropriate personal protection behaviors can effectively reduce individual exposures to tick bites [14]. Unfortunately, basic knowledge of ticks and preventive behaviors are often limited, even among public health professionals, and locally targeted surveillance programs to inform risk may be lacking [15–19]. To address these gaps, efforts to improve general knowledge of ticks and TBDs in the United States have been spearheaded by the Centers for Disease Control and Prevention (CDC). These include educational workshops, online and printed guides to TBDs written for healthcare professionals, supporting the expansion of state-level tick surveillance efforts, and promoting research innovations through several regional Centers of Excellence (COEs) in Vector-Borne Diseases [18, 20]. In 2020, the Southeastern Center of Excellence in Vector-Borne Diseases (SECOEVBD) began developing an Online Tick Training Course (OTTC), comprising a series of educational modules on ticks, tick management, and TBD risk and prevention. While openly accessible, these materials were expressly developed to provide foundational knowledge on ticks and TBDs for public health and vector control professionals. In order to ensure that the course was both useful in increasing knowledge, and to assess the utility of that knowledge gain for participants, evaluation and assessment components were explicitly included in its development. Here, we summarize the outcomes from the first round of assessment (May 2021-December 2022) of the OTTC.

## Methods

### Online training course

The Online Tick Training Course (OTTC) was developed cooperatively by the SECOEVBD, University of Florida, and Old Dominion University. The course consists of seven online modules, delivered through the UF Institute of Food and Agricultural Sciences (UF IFAS) Extension Canvas interface. The modules cover a variety of topics, including tick biology and identification, tick surveillance and control, tick-borne diseases, public health, and tick personal protective behaviors and bite prevention.

While the OTTC is freely available to the public, the course was promoted through vector control and public health agencies throughout the Southeastern United States. To incentivize the participation of active public health and vector control employees, participants who completed the OTTC were eligible to receive Continuing Education Units (CEUs), which are needed to maintain professional certifications with the Florida Department of Agriculture and Consumer Services (FDACS) and the California Department of Public Health or other state organizations with prior approval.

### Course evaluations

The study protocol and survey tools were reviewed and approved by the Institutional Review Board (IRB) at the University of Florida (IRB 202100687). Participants who enrolled in the OTTC were recruited for this study and were given informed consent materials and the option to opt-in the course assessment study prior to starting online training modules. Four survey tools were used to collect data on participant outcomes to evaluate the impact of the OTTC. These included a pre-course assessment, a post-course assessment, an immediate post-course student evaluation survey, and a delayed course evaluation survey. The pre-course and post-course assessment tools consisted of nine questions, related to topics covered in the OTTC (tick biology and identification, n=4; tick-borne diseases, n=2; safe tick removal, n=1; and public health and surveillance, n=2). The pre-course assessment was administered after informed consent was obtained, and before participants started any training materials, and the post-course assessment was delivered to participants immediately after successful completion of the OTTC. The post-course student survey was also delivered to participants at the end of the OTTC, and featured questions on perceived knowledge gained, intent to use knowledge, and facilitators and barriers to use (S2 Table). Delayed course assessments were administered approximately six months after successful completion of the OTTC and consisted of questions on how participants applied course content in their occupation, elements of the course participants found useful, barriers to applying course content, and open text fields for users to expand on answers. Participants who opted-in were contacted via email and provided a link to the assessment tool via the Qualtrics platform (https://www.qualtrics.com, Provo, UT, USA).

Survey responses for respondents were recorded and summarized to show trends in baseline knowledge, the immediate impact of the OTTC, and long-term utility and retention of knowledge from the course. We used a paired sample sign test to assess statistical differences in pre-course and post-course testing outcomes [21]. All statistical analyses were performed in R (ver. 4.1.2), and data visualizations were conducted in R and ArcGIS Pro (ver. 3.1.0).

## Results

Participation in the OTTC – A total of 457 participants signed up to take the OTTC in the program’s initial launch (May 2021 through December 2022). There were 319 of these participants who opted into the OTTC assessment study and of these, 317 (99.4%) completed the pre-course assessment. A total of 255 (79.9%) participants who opted into the study also completed the post-course assessment and successfully finished the OTTC. Participants were primarily located throughout the Southeastern United States. While OTTC training was developed and promoted for stakeholders in the Southeastern US, we also found high user engagement throughout the continental US, particularly in California (Fig. 1).

**Fig. 1.**
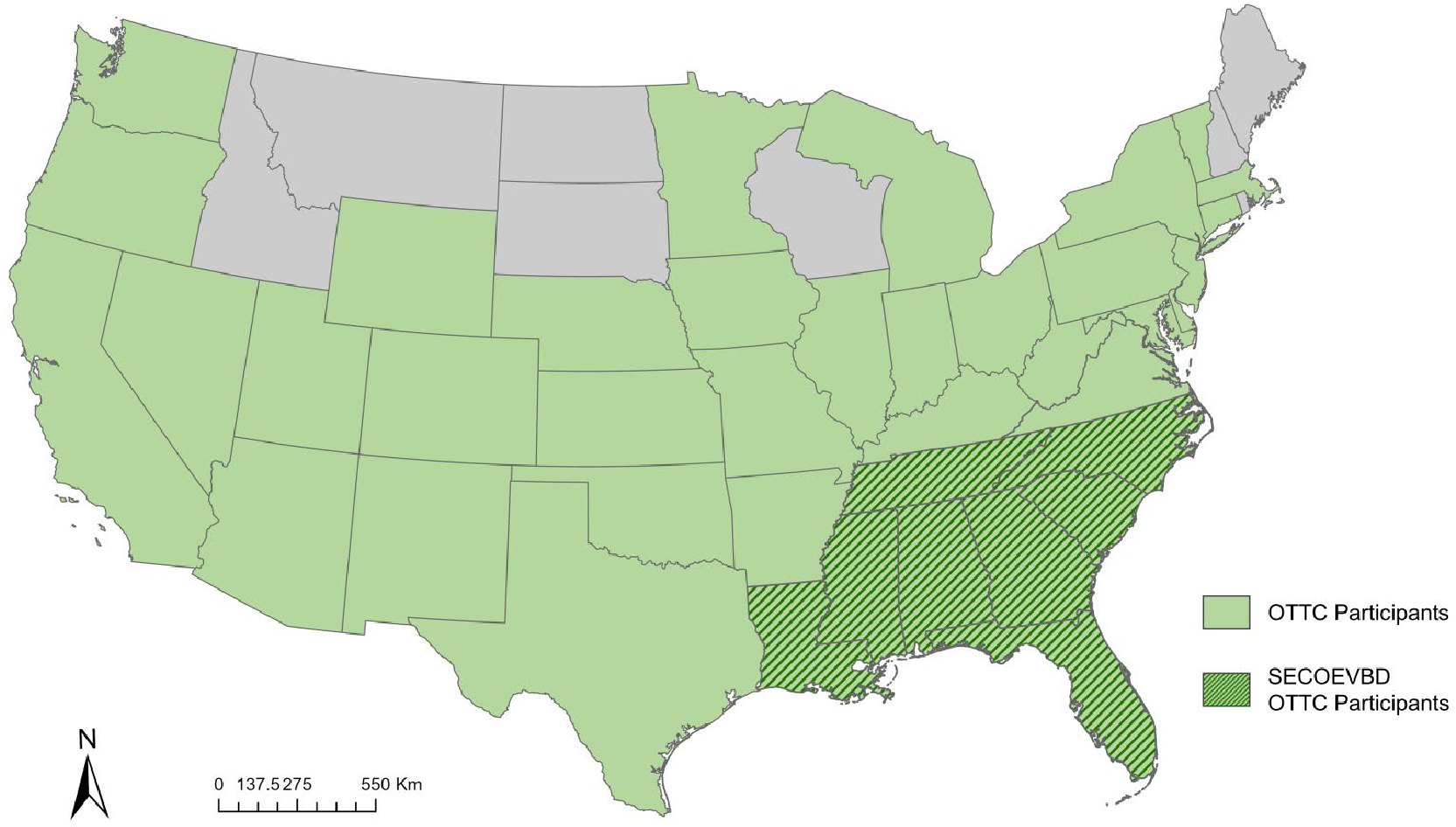
States in the continental US where OTTC participants were located. Dark green hash indicates states represented by the Southeastern Center of Excellence in Vector-Borne Diseases (SECOEVBD) at the time of the OTTC launch.

There was also a small number of international OTTC participants who accessed the course from countries including Australia, Brazil, Canada, Ecuador, India, Japan, Mexico, Nigeria, Pakistan, and the United Kingdom.

Participants in the OTTC represented a variety of professional roles within pest management and vector control. The majority (66.4%) of enrollees indicated that the OTTC was either “very” or “extremely” relevant to their current line of work. Many participants in the OTTC (37.2%) indicated that they worked in vector control. Under one quarter (20.2%) of all vector control professionals indicated that they were certified operators and 7.5% were supervisors of technicians. Over one quarter (27.7%) of participants self-identified as working in the public health field, which included state health department employees, epidemiologists, environmental scientists, and public health entomologists. Over one third (38.3%) of participants indicated that they were directly involved in tick surveillance or control in some capacity, and 13.8% focused on human surveillance of TBDs. Many respondents (28.5%) answered that their working unit or agency was not directly involved or connected with tick control or surveillance, where some noted that while ticks were of interest, resources were predominantly dedicated to mosquito control. Many participants were also connected with academic institutions, either through involvement in research activities (8.7%), or as students (14.6%). Fields with relatively lower representation included military (2.8%), veterinary surveillance and agricultural agencies (0.8%), and medical providers and clinicians (0.4%). A small percentage (1.2%) of participants were not professionally associated with public health, vector control, or research, but rather, were members of the general public who wanted to learn more about ticks, or who were concerned about tick activity in their communities or on their property.

### Pre-Course and Post-Course Assessment

The pre-course assessment survey was used to establish baseline knowledge of participants on tick biology and control before accessing OTTC materials. The majority of participants who opted into the study completed the pre-course assessment (n=317). The mean pre-course test score for participants who completed the course (n=255) was 62.6%, with most participants (> 50%) incorrectly answering questions on larval and nymphal tick characteristics, and how to determine the sex of ticks. The majority of participants (94.5%) were able to correctly identify the best method for tick removal (i.e., using fine-tipped tweezers or a tick removal tool) before taking the OTTC. Participants also had fair knowledge of early TBD symptoms (75.6%), efficacy of control methods across tick species and life stages (85.9%), and the general lack of tick surveillance data for public health (82.0%). The majority of participants (79.9%) completed the OTTC and post-course assessment (n=255). Testing outcomes improved significantly upon completion of the OTTC, where the average post-course assessment score was 86.7%, and paired scores for each question were significantly higher (Table 1, Fig. 2).

**Table 1.** Questions (Q) and paired outcomes for the OTTC pre-course (PRE, n=255) and post-<u>course (POST, n=255) assessment.</u>

| Q | PRE | POST | p-value |
| --- | --- | --- | --- |
| 1 Larvae have (select all that apply) | 49.8% | 77.30% | < 0.0001 |
| 2 You can tell the difference between male and female nymphs with a microscope (T/F) | 38.8% | 77.30% | < 0.0001 |
| 3 To determine the sex and life stage of the tick (select all that apply) | 34.2% | 59.60% | < 0.0001 |
| 4 Which is true regarding Lyme disease? | 54.1% | 97.60% | < 0.0001 |
| 5 That best way to remove a tick from yourself is | 94.5% | 100% | < 0.0001 |
| 6 Ticks find hosts through which of the following (select all that apply) | 48.8% | 82.90% | < 0.0001 |
| 7 Common symptoms of the early stages of nearly all tick-borne diseases include (select all that apply) | 75.6% | 92.40% | < 0.0001 |
| 8 All tick control methods have the same efficacy on all tick species and life stages (T/F) | 85.9% | 99.20% | < 0.0001 |
| 9 There is a lack of tick data because fewer than half of public health entities fund any work related to tick surveillance (T/F) | 82.0% | 95.30% | < 0.0001 |

**Fig. 2.**
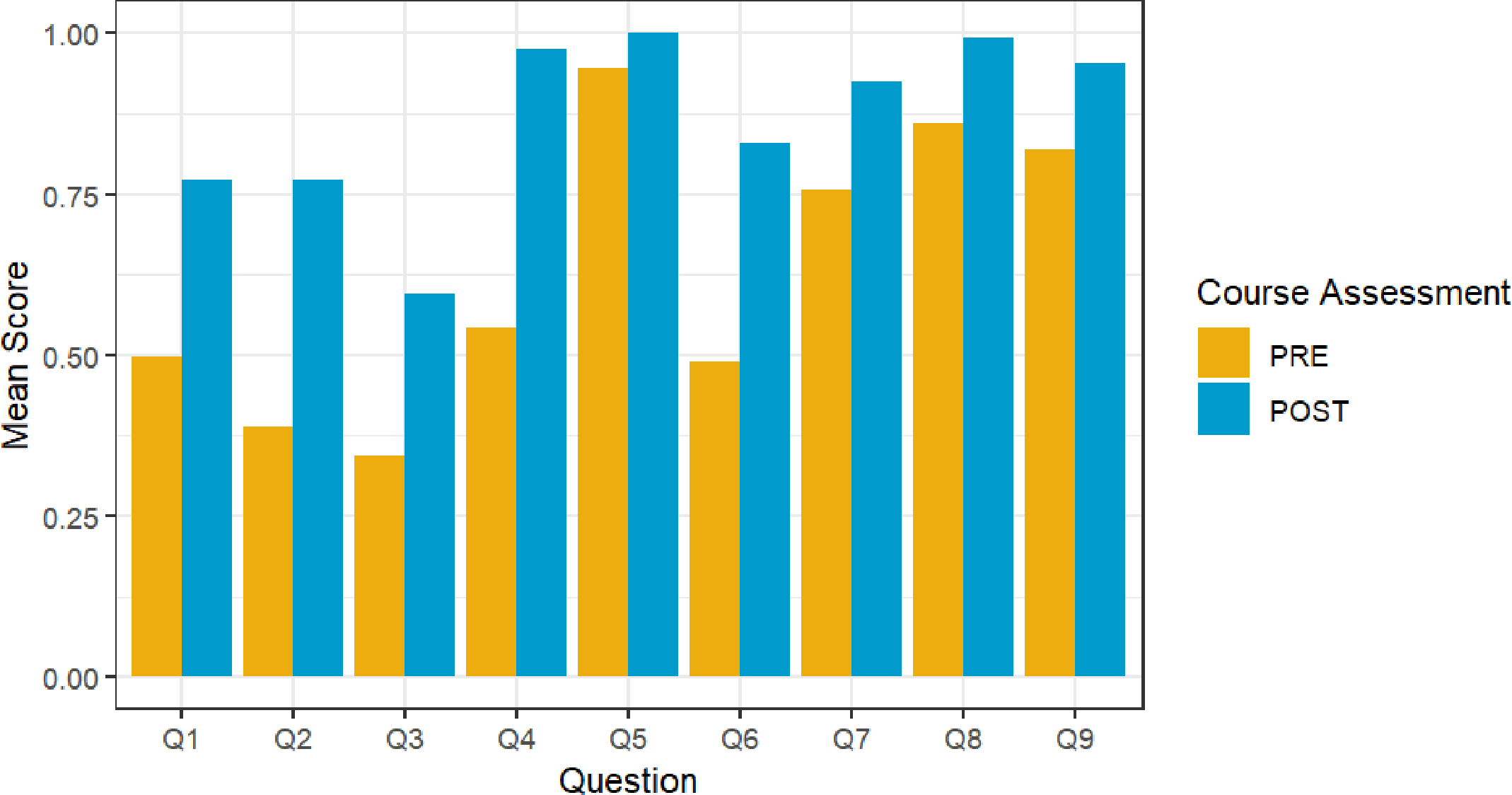
Comparison of average paired scores on questions from participants (n=255) who completed both the pre-course (orange) and post-course (blue) OTTC assessments.

While there was no single question that the majority of participants answered incorrectly in the post-course survey, the lowest scoring question (59.6%) was a multiple answer format knowledge question (i.e., “select all that apply”) on ways to determine the sex and life stage of ticks. Participants scored very high (>92%) on five questions in the survey, including questions on Lyme disease risk, tick removal, TBD symptoms, tick control methods, and lack of tick surveillance data. The largest gains in knowledge (i.e., pre-course versus post-course scoring) were seen on questions related to tick biology and identification (Q1-Q3, Q6), and one question on Lyme disease (Q4) (Fig. 2).

### Student Survey of Course Materials

At the conclusion of the course, most participants (63.6%) felt that they would definitely use what they learned in the OTTC in their work. Overwhelmingly (97.6%), participants felt that the course had an appropriate balance of lectures and interactive training materials, and most participants (62.8%) had no suggestions for further improvement to the OTTC. In open text responses, the use of images, videos, and interactive quizzes were consistently identified as successful elements that facilitated learning and comprehension of content. Some suggestions for course improvement repeatedly mentioned in open text responses included the addition of printable materials to summarize and review course content and printable booklets on tick identification for use in the field. Inclusion of a module on molecular techniques and diagnostic tools for TBDs was also suggested as a potential addition to future iterations of the training.

### Delayed Course Assessment

Fifty-four (16.9%) participants who completed the OTTC also completed the delayed course assessment. Eighteen (33.3%) participants indicated that the information learned in the OTTC was used frequently in their work, while the majority (59.3%, n=32) of participants responded that the training was used in their work to some extent. Few participants (7.4%) did not use information from the OTTC in their work at all. Participants were asked to identify which information from the OTTC was most used in their jobs. While responses varied, tick biology and ecology (40.7%), tick identification (33.3%), public health surveillance (22.2%), and TBD transmission (7.4%) were commonly identified as the most useful applied information. A small number of individuals (2.1%) additionally noted that they were leveraging information learned in the OTTC to help develop and improve content for university courses or agency outreach programs. The majority also stated that having opportunities to apply what was learned (70.4%), having time to apply course concepts (53.7%), and having reminders of key concepts and skills (50%) were major factors that facilitated the use of course information and concepts in their work. Having necessary resources (42.6%) and the support of colleagues (33.3%) or supervisors (29.6%) were also identified as important factors that helped participants apply course knowledge on the job.

Participants were also asked to identify major barriers to using OTTC content in real-world settings. Chief among these were lack of opportunities to apply skills and concepts at work (18.5%) and the need for specialized training in the subject matter beyond what was covered in the course (18.5%). Several respondents answered that course content was not relevant to their work at all (14.8%), or that they did not remember concepts well enough to apply them at work (9.3%). Other barriers to applying content provided in open text responses included lack of an existing agency tick control program, the end of the vector control season, and modules that were too broad for specialized needs. Two participants suggested that access to copies or summaries of course information would provide a useful reference to use in their job. One participant requested more content that could be applied by physicians and healthcare providers in clinical settings, such as symptoms and available diagnostic tests for different TBDs.

## Discussion

Surveillance and control efforts are cornerstones of vector-borne disease management programs in the United States. Yet, despite the prevalence and rising burden of TBDs, many public health vector control programs predominantly focus surveillance efforts and resources on medically important mosquitoes [18, 19]. Resources are increasingly dedicated for research and surveillance efforts for TBDs, which include pipelines for agency support, the development of open access data platforms (e.g., the VectorByte data hub) to promote increased sharing and accessibility of tick surveillance data [22], and indeed, the very creation of the SECOEVBD (17). Nevertheless, gaps in baseline knowledge and training regarding ticks and TBD risk, particularly among public health and vector control practitioners, may contribute to inconsistent tick surveillance and control efforts, exacerbating issues such as underreporting of cases, lags in detection of expanding vector populations, or insufficient public outreach and promotion of established preventive behaviors. The SECOEVBD designed the OTTC to address these gaps, providing a freely accessible online course to provide baseline knowledge on ticks, TBDs, and their management. Although the OTTC is currently available to anyone with an interest in ticks, the course was primarily advertised to public health and vector control professionals prior to its initial launch in 2021 (platform migration is underway at the time of this writing, 2024). This effort was reflected in our initial survey results, where the majority of participants self-identified as either vector control professionals or public health department employees. The OTTC was effective in increasing participants’ knowledge across all subject areas, including tick biology, identification, diseases and safety, and public health and surveillance.

The OTTC was well received, with the majority of participants deeming the training important to their work. Participants largely felt that the format of the course struck a good balance between lectures and interactive materials, and most felt there was no room for further improvement. However, several changes were suggested that may improve the utility of the course for users, while being potentially easy to implement. These included production of summarizations (i.e., “fact sheets”) for material review and reference after conclusion of the course, and the addition of training subjects that could be useful to epidemiologists and clinicians, namely, a review of molecular surveillance methods and diagnostic tools for TBDs in humans. Despite the overall positive reception of the OTTC, participants identified several barriers that prevented them from using knowledge gained in an applied capacity, specifically citing either a need for more specialized information (e.g., clinicians and veterinary pathologists interested in clinical symptoms and diagnostic tools), or a lack of opportunities to apply what they learned (e.g., vector control operators working in districts with no established tick surveillance or control programs).

The use of CEUs may have incentivized participation in the OTTC, particularly for public health vector controllers who need to maintain professional certifications for their jobs, for example, certified pesticide applicators. Although CEUs were developed to meet the requirements for FDACS certification, anyone could take the course online and request approval for CEUs to apply to certifications through other states and agencies. The CEU incentive likely underlies the geographic diversity in participation observed, where many people enrolled in the course were not located in the southeastern US. Offering CEUs may have helped to bolster the number of participants, but may have also unintentionally skewed some of our post-course and delayed assessment responses, in particular, questions relating to applied use of OTTC knowledge. For example, some noted that the tick species covered in the course did not include some medically important vectors in their area. One participant reported via an open text response that they were not able to apply course knowledge, because they worked in urban vector control in a metropolitan area beyond the SECOEVBD purview, and had only encountered a single tick in over 15 years on the job.

There was considerable attrition in the delayed evaluation, compared to the post-course assessment taken at the end of the OTTC. Only a small fraction of participants responded to the delayed evaluation prompt, which was sent via email approximately six months after course completion. While the delayed assessment provided useful insights on how participants were applying the OTTC training at their jobs, in practice, the timeline for follow-up may be too long. In future iterations of the OTTC, shortening the time between course completion and delayed assessment may help increase the number of responses. The delayed assessment was also administered via the Qualtrics platform, while the immediate post-course assessment surveys were administered through Canvas, the same interface as the OTTC. Adoption of a single, consistent survey platform may also help improve participation in the delayed assessment.

The results of the OTTC assessment and surveys provided critical insights into the efficacy of training materials, as well as participants’ occupation and perceptions surrounding the course’s value, and barriers to applying knowledge in their fields. While the course was successful in its primary goal of increasing participant knowledge, through the surveys we have also identified areas for improvement that can be incorporated into future iterations of the OTTC. Questions in survey tools could be expanded to capture additional, or more nuanced information, such as motivations for taking the course (e.g. CEUs), geographic locations where knowledge will be applied, existence of tick surveillance programs at their job, and more. Further, categories of answers could be improved based on commonalities observed in open text survey responses. For example, in the question on occupation, many respondents indicated that they were academic researchers or students, but these categories were not among selectable options (i.e., users would have to respond with ‘Other’ and manually record their occupation in an open text field).

Although the course was primarily designed for, and promoted to, vector control and health department employees, we saw some diversity in the occupations of participants. Identifying cross-disciplinary channels of dissemination to promote the OTTC may help reach other professional communities that would benefit from comprehensive tick training. These include vector-borne disease researchers in academia, medical entomology and public health students, practicing clinicians, and employees with agricultural agencies tasked with disease research and veterinary surveillance. While some respondents indicated a need for more specialized training in tick-borne diseases and surveillance, or development of modules for other geographic areas, this is currently beyond the scope of the OTTC, which was intentionally designed to establish baseline knowledge for professionals in the southeastern US. Nevertheless, with sufficient interest, the OTTC could serve as a model for the development of future online trainings to meet the needs of specific professional groups.

## Conclusions

Tick-borne diseases have been garnering increased attention over the past decade in the United States due to increasing burden and expanding geographic distributions. Vector control districts and public health agencies are often tasked with surveillance and control activities for ticks and the diseases caused by the pathogens they transmit, yet, baseline knowledge on ticks, disease risk, and management may be lacking. In this study, we demonstrated that the OTTC developed by the SECOEVBD has successfully addressed this gap, significantly improving knowledge of ticks and tick-borne disease risk in key professional groups. Information on tick biology, ecology, and identification showed some of the biggest improvements in knowledge, and these subjects were identified as most used on the job in the delayed assessment.

## Data Availability

All data produced in the present study are available upon reasonable request to the authors

## List of Abbreviations

CDC: Centers for Disease Control
CEU: Continuing Education Units
FDACS: Florida Department of Agriculture and Consumer Services
OTTC: Online Tick Training Course
SECOEVBD: Southeastern Center of Excellence in Vector-Borne Diseases
TBD: tick-borne disease
UF IFAS: University of Florida Institute of Food and Agricultural Sciences

## Declarations

### Ethics approval and consent to participate

The study protocol and survey tools were reviewed and approved by the Institutional Review Board (IRB) at the University of Florida (IRB 202100687). Participants were recruited through the OTTC, and were given informed consent and the option to opt-in the course assessment study prior to starting online training modules.

### Consent for publication

Not applicable

### Availability of data and materials

De-identified data can be requested directly from the authors, subject to the terms of the IRB as approved

### Authors’ contributions

SJR and HDG conceptualized the study. SJR, HDG, and ALW developed online training course materials and survey tools. CAL collected and prepared data for analysis. CAL and SJR performed statistical analyses and interpreted data. CAL, SJR, HDG, and ALW drafted the manuscript. All authors read and approved the final manuscript.

## Acknowledgements

The authors would like to thank Kaci McCoy and UF IFAS for assistance in the production of this course, and Dr. Heather Coatsworth for assistance with surveys and data collection. Additionally, we would like to thank the partners of SECOEVBD who provided critical feedback before launching the OTTC.

## Supplemental Materials

**S1 Table.** Questions and outcomes for the OTTC pre-course and post-course assessment, applying an unpaired two-sample Wilcoxon test on mean testing outcomes from the pre-course <u>and post-course assessments.</u>

| Question |  | PRE<br>(n=317) | POST<br>(n=255) | p-value |
| --- | --- | --- | --- | --- |
| Q1 | Larvae have (select all that apply) | 50.10% | 77.30% | < 0.0001 |
| Q2 | You can tell the difference between male and female nymphs with a microscope (T/F) | 42.00% | 77.30% | < 0.0001 |
| Q3 | To determine the sex and life stage of the tick (select all that apply) | 34% | 59.60% | < 0.0001 |
| Q4 | Which is true regarding Lyme disease? | 52.70% | 97.60% | < 0.0001 |
| Q5 | That best way to remove a tick from yourself is | 95.00% | 100% | 0.0003 |
| Q6 | Ticks find hosts through which of the following (select all that apply) | 50.60% | 82.90% | < 0.0001 |
| Q7 | Common symptoms of the early stages of nearly all tick-borne diseases include (select all that apply) | 77.00% | 92.40% | < 0.0001 |
| Q8 | All tick control methods have the same efficacy on all tick species and life stages (T/F) | 87.70% | 99.20% | < 0.0001 |
| Q9 | There is a lack of tick data because fewer than half of public health entities fund any work related to tick surveillance (T/F) | 83.60% | 95.30% | < 0.0001 |

**S2 Table.**
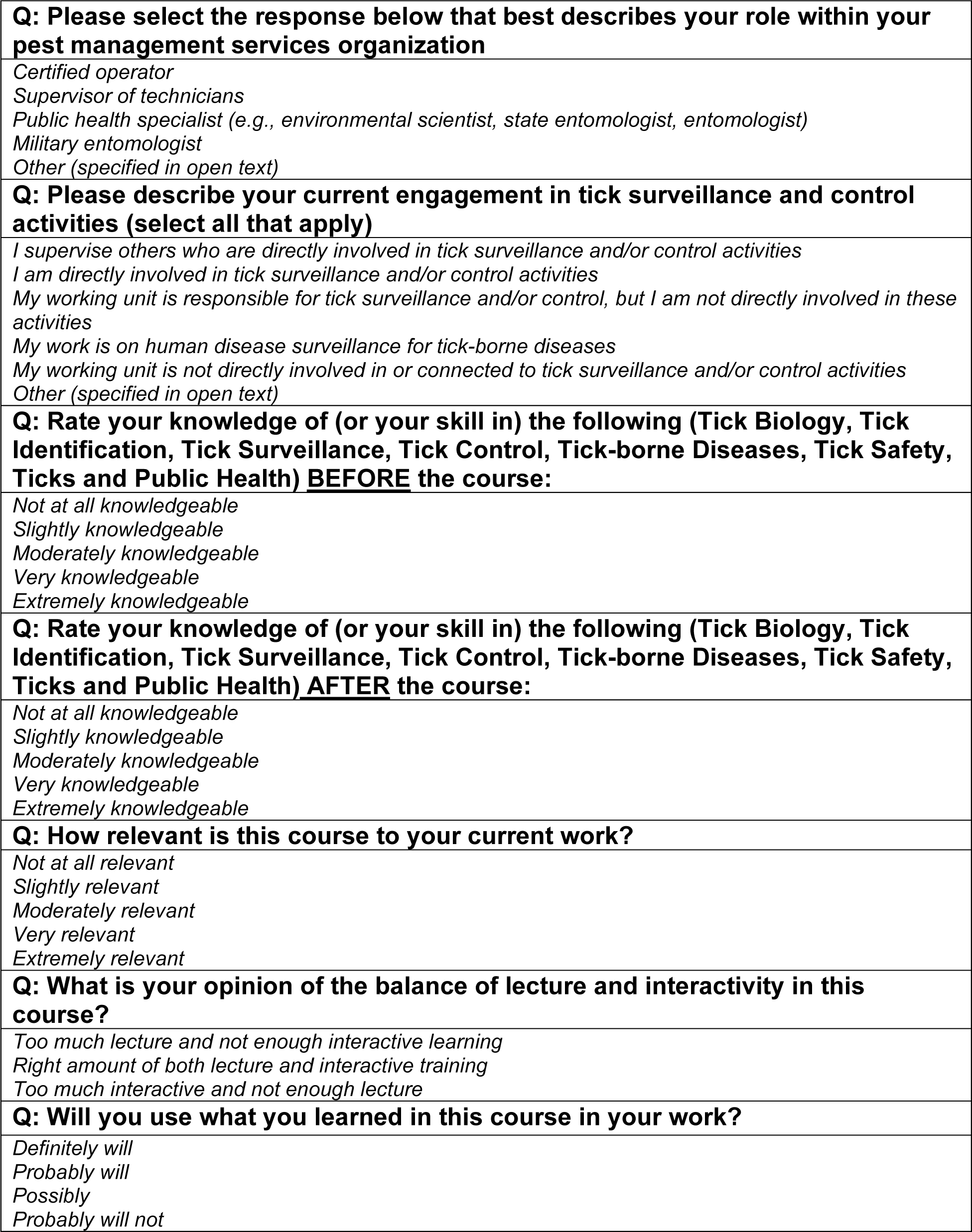
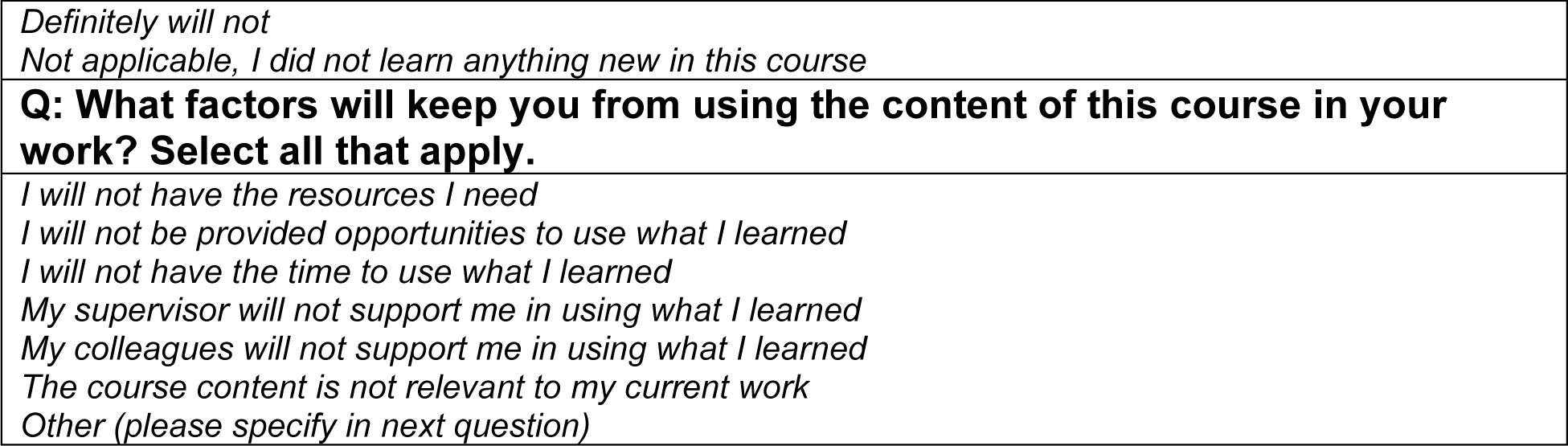
Questions and multiple-choice answer choices included in the post-course student assessment. In addition to these questions, the full assessment also included several sections for students to provide feedback via open text responses.

**S2 Fig.**
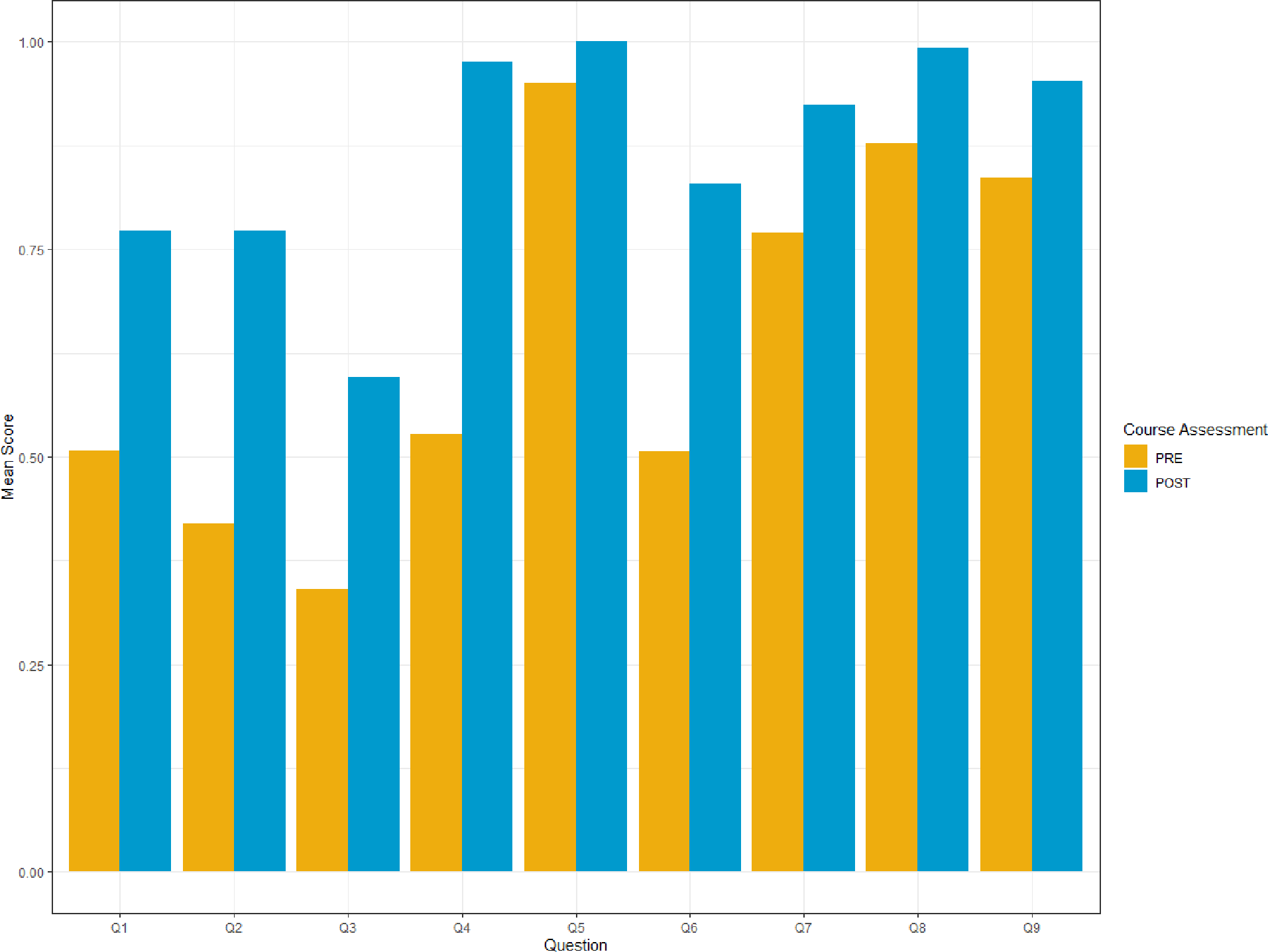
Comparison of average scores on questions from all participants who completed the pre-course (n=317, orange) and post-course (n=255, blue) OTTC assessments.

